# Estimation of protection for COVID-19 in children from epidemiological information and estimate effect of policy in Japan

**DOI:** 10.1101/2020.03.27.20045252

**Authors:** Junko Kurita, Yoshiyuki Sugishita, Tamie Sugawara, Yasushi Ohkusa

## Abstract

**Background:** Incidence in children was much less than in adults during the COVID-19 outbreak. Sports and entertainment events were canceled (VEC) in Japan for two weeks during 26 February – 13 March. Most schools were closed (SC).

**Object:** We construct a susceptible–infected–recovered model using three age classes and estimate the basic reproduction number (R_0_) and protection level among children simultaneously. Then we simulate SC and VEC effects.

**Method:** We used data of patients with symptoms in Japan during 14 January to assess SC and VEC introduction. Effects of SC and VEC were incorporated into the model through change in the contact pattern or frequencies among age classes.

**Results:** Results suggest R_0_ as 2.86 [95%CI of 2.73, 2.97]. The protection level was estimated as 0.4 [0.2, 0.7]. SC and VEC can reduce the total number of patients significantly, by 6–7%.

**Discussion and Conclusion:** The estimated R_0_ was similar to that found from other studies in China and Japan. We found a significant protection level among children, and by effects of SC and VEC. Introduction

## Introduction

The initial case of COVID-19 in Japan was that of a patient who showed symptoms when returning from Wuhan, China on 3 January, 2020. As of 18 March, 2020, 690 cases were announced as infected in the community, including asymptomatic cases, but excluding those abroad in countries such as China or those infected on a large cruise ship: the Diamond Princess [1].

Earlier research suggested that children might be less susceptible to COVID-19 for lifestyle-related factors such as lack of smoking and less vigorous immune response than that of adults [2]. Children might have some cross protection from non-novel coronavirus infection. Even though the reasons for it had not become clear, the number of children among patients was quite small. In fact, even in Japan, the incidence rate including asymptomatic cases among children was 0.79 per million population compared to 5.97 for adults and 7.82 for elderly people as of 17 March. Incidence among children was approximately one-tenth that of elderly people. Therefore, this clear phenomenon must be incorporated into the model when predicting the outbreak. On that point, we produce a susceptible–infected–recovered (SIR) model with three age classes and search for a protection rate in children as well as the basic production number, R_0_.

In Japan, school closure (SC) and voluntary event cancellation (VEC) were introduced on 27 February. The former was intended to decrease contacts among children. The latter was aimed at decreasing contact among adults. We also examine simulation of these policies as hypothetically changing contact patterns and frequencies.

## Method

We applied a simple SIR model [3] with three age classes: children 19 years old or younger, adults younger than 59 but older than 20 years old, and elderly people 60 years old or older. The incubation period was assumed to be equal for three age classes and following the empirical distribution in the outbreak in Japan.

Experiences of Japanese people living in Wuhan until the outbreak provide information related to mild cases because complete laboratory surveillance was conducted for them. During January 29 – February 17, 2020, 829 Japanese people returned to Japan from Wuhan. All had received a test to detect COVID-19; of them, 14 were found to be positive for COVID-19 [4]. Of those 14, 10 Japanese people had exhibited mild symptoms; the other 4 showed no symptom. Moreover, two Japanese residents of Wuhan exhibited severe symptoms: one was confirmed as having contracted COVID-19. The other died, although no fatal case was confirmed as COVID-19 by testing. In addition, two Japanese residents of Wuhan with mild symptoms were refused re-entry to Japan even though they had not been confirmed as infected. If one assumes that the Japanese fatal case in Wuhan and that the two rejected re-entrants were infected with COVID-19, then 2 severe cases, 12 mild cases, and 4 asymptomatic cases were found to exist among these Japanese residents of Wuhan. We therefore apply these proportions of asymptomatic cases to symptomatic cases in the simulation.

Assuming that the power of infectivity among severe patients and mild patients are equal among asymptomatic cases, half of the symptomatic cases can be assumed. This assumption about relative infectiousness among asymptomatic cases compared with symptomatic cases was also assumed in simulation studies for influenza [5–9].

We presumed some fraction of children were immune. No information about it was available ex ante: we searched it in the range of (0,1). Simultaneously, we sought R_0_ to fit the number of patients during 14 January – 17 March and to minimize the sum of absolute values of discrepancies among the reported numbers and the fitted values. Its 95% confidence interval (CI) was calculated using 10,000 iterations of bootstrapping for empirical distribution.

The contact patterns among children, adults, and elderly people were estimated in an earlier study [10]. We identified contact patterns as follows: the share of children contacting with other children was 15/19 of all contacts, with adults was 3/19, and with elderly people was 1/19; the share of adult contacting with children was 3/9 of all contacts, with other adults was 5/9, and with elderly people was 1/9; the share of elderly people contacting with children was 1/7, with adults was 2/7, and with other elderly people was 4/7.

Experience in Japan showed the pneumonia incidence in elderly COVID-19 patients as 30.6%. That among adults was 22.2%. We assumed these numbers as a proportion of severe cases to symptomatic cases. Among children, no pneumonia case was reported. Only pneumonia cases were administered. Its length of hospitalization was assumed as 30 days. Of those, 30% of severe cases were assumed to use an intensive care unit and respirator.

We used data of the community outbreak of patients with COVID-19 who showed any symptom in Japan for 14 January – 28 February, 2020. From 2 March, all schools were closed down as social mitigation for COVID-19. Contact patterns among students or between students and adult were thereby altered considerably. Therefore, we do not use data for the period during which schools were closed ad hoc for estimation. We excluded some patients who had returned from abroad, such as from China, and those who were presumed to be infected persons from the Diamond Princess. They were presumed to be not community-acquired in Japan.

Published information about COVID-19 patients with symptoms from the Ministry of Labour, Health and Welfare (MLHW) Japan was usually affected adversely by some delay because of uncertainty during onset to visiting a doctor or in the timing of a physician’s suspicion of COVID-19. Therefore, published data of patients must be adjusted at least a few days. To adjust the data, we applied the following regression. We denote *Xt-k*|*t* as the number of patients for whom the onset date was *t-k* published on day *t*. The dependent variables are the degree of reporting delay, *Xt-k-m*|*t* / *Xt-k-m*|*t-m*, where *k*>*m* in several *m* and *k*. Here, *m* denotes the difference of the publishing dates between the two published. Date *t* represents the publishing date of the latest publishing. The explanatory variables were 1/*k*, 1/*m*, and 1/*km*. The degree of reporting delay was estimated as [estimated coefficient of constant term] + [estimated coefficient of 1/*k*]/*k*, when *m* was sufficiently large and time had passed. Therefore, this estimated degree of reporting delay multiplied by the latest published data is expected to be a prediction of the number of patients for whom the onset date was *t*-*k*. We used this adjusted number of patients in the latest few days including those after VEC was adopted. We used published data of 2, 5, 6 and 9–17 March, 2020 provided by MLHW [1].

First, we estimated the joint distribution of R_0_ and the protection rate in children in Japan to fit the data of community outbreak before SC and VSC introduce on 27 February. Using the estimated R_0_ and protection rate in children, we simulate the effect of SC and VSC assuming hypothetical change in the contact matrix. We assumed that the SC decrease contact frequencies among children to be one-tenth, the rise in contacts of children with adult to be twice, the rise in contacts of adults with children to be four-thirds by 33.3%, and the decline in contacts among adults by 20%. Conversely, VEC was assumed to decrease contacts among adults by 20% and among elderly people by 25%. In the simulation with application of both of SC and VEC, the contact matrix was assumed to change to be the sum of effect of SC and VEC. To evaluate SC and VEC, we compare the total number of patients with any symptoms between the base case and intervention. We calculate 95% CI through bootstrapped distribution of total number of patients with intervention over the total number of patients in the base case.

## Ethical consideration

All information used in the present study has been published previously. There is therefore no ethical issue.

## Results

During 14 January – 28 February in Japan, 9 cases in children, 165 cases in adults, and 126 cases in elderly people were identified as community-acquired COVID-19 for whom the onset date was published. Figure 1 depicts the empirical distribution of incubation period among 62 cases for which the exposed date and onset date were published by MHLW. Its mode and median were six days; the average was 6.74 days.

**Figure 1:**
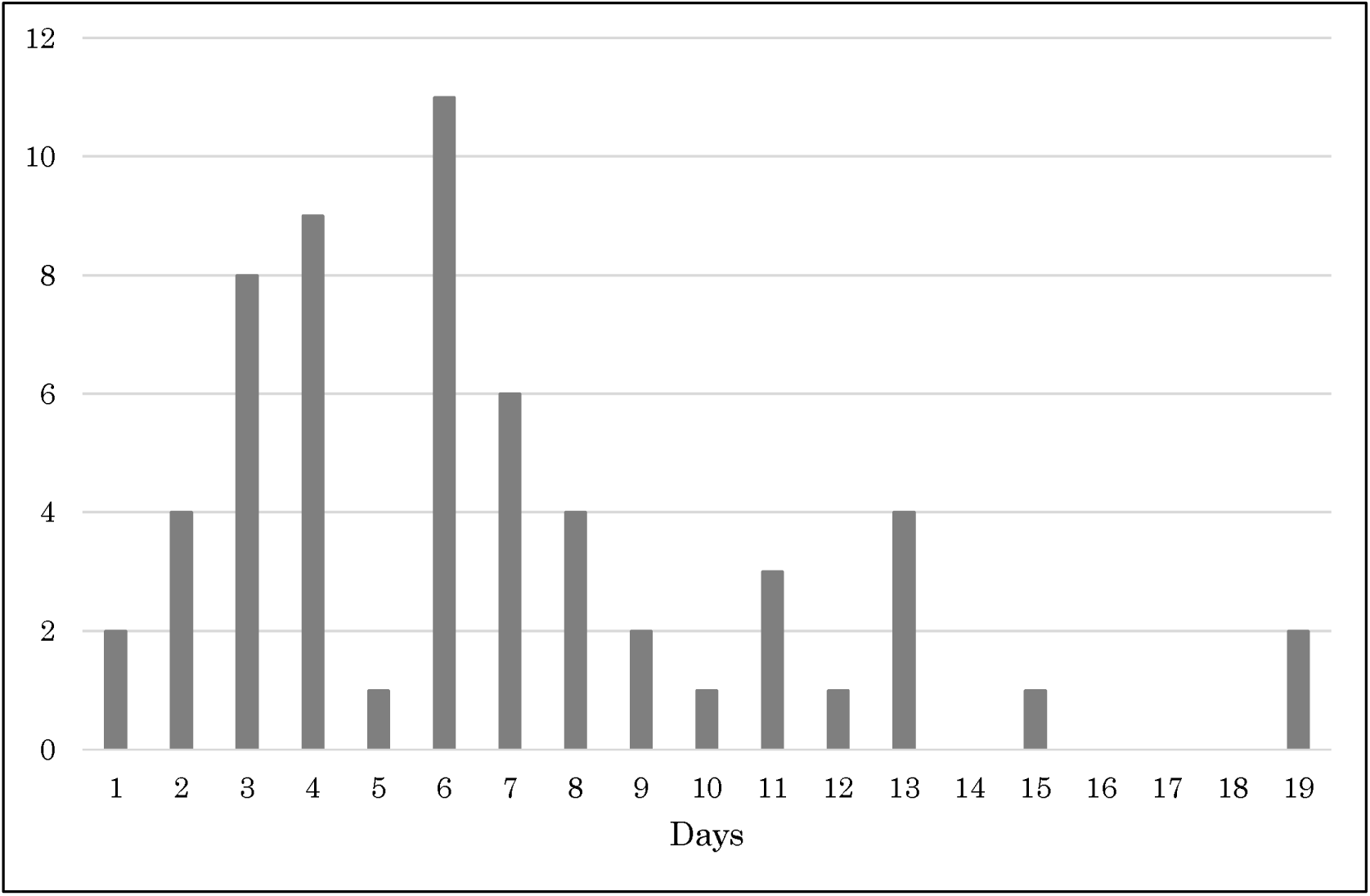
Empirical distribution of incubation period published by Ministry of Labour, Health and Welfare, Japan (number of patients) Notes: Bars represent the number of patients by incubation period among 62 cases whose exposure date and onset date were published by Ministry of Labour, Health and Welfare, Japan.

Figure 2 depicts epidemic curves published at 2, 5, 6, 10, 11, 12, 13, and 14 March. From this information, we estimated the degree of reporting delay. Those results are presented in Table 1. The table shows that 1*/k*, 1/*m*, and 1/*km* are all significant. When *m* is sufficiently large, the effects of 1/*m* and 1/*km* converge to zero. Therefore, the estimated degree of reporting delay consists of the term of 1/*k* and a constant term. Based on these results, we predict the degrees of reporting delay as 19.3 for *k*=1, 9.64 for *k*=2, 6.42 for *k*=3, and so on.

**Table 1:**
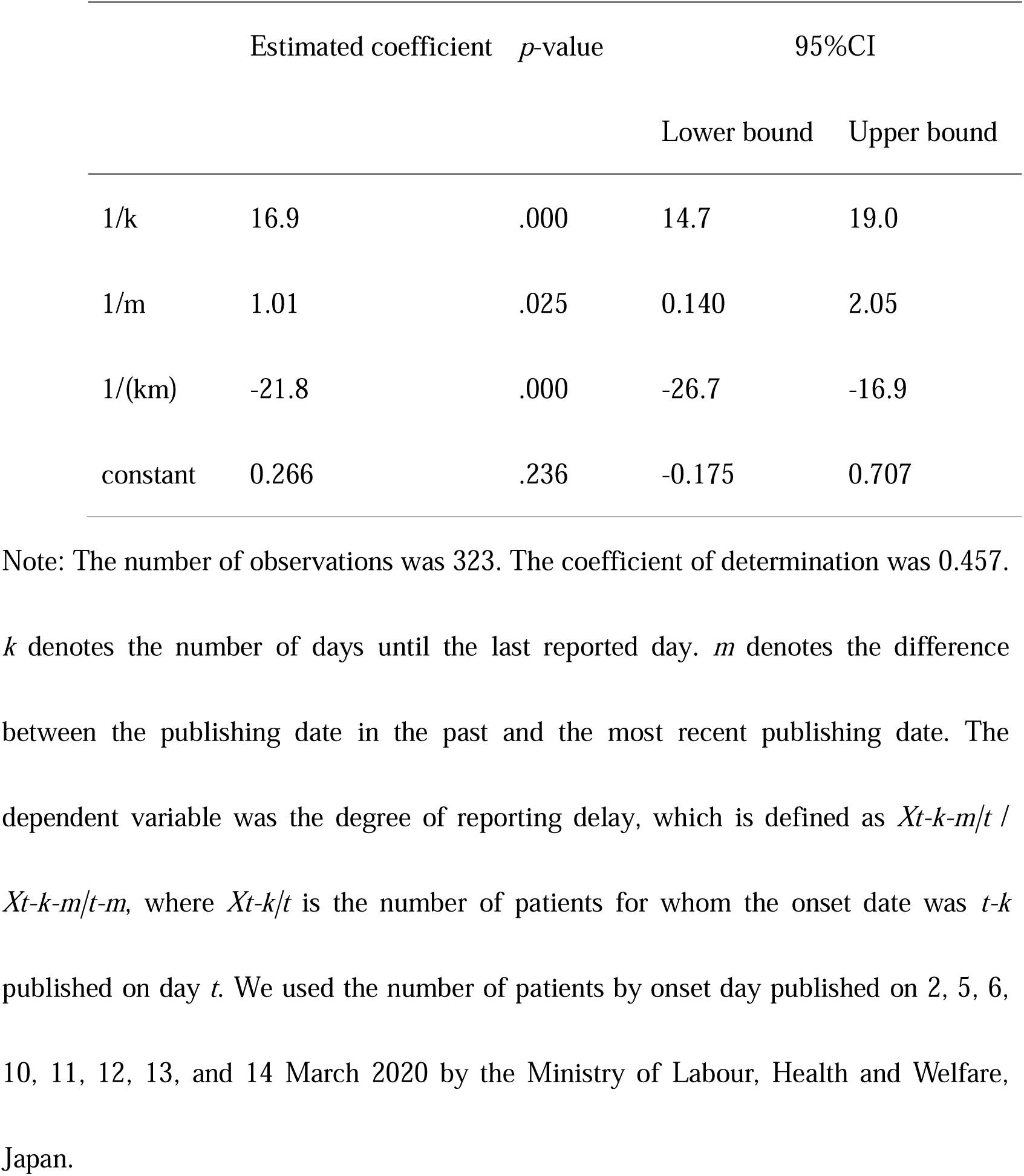
Estimation results of the degree of reporting delay

The R_0_ values were estimated as 2.86 [95%CI 2.73, 2.97]. The protection level was estimated as 0.4 [95% CI 0.2, 0.7].

In the base case, presented as Table 1, the total number of patients with any symptom was estimated as 78.4 [69.5, 83.3] million. The maximum number of patients with new onset was estimated as 2.25 [1.85, 2.58] million per day at the peak.

At the peak, the maximum number of the administered patients with pneumonia was estimated as 12.9 [11.7, 13.8] million per day. Moreover, the maximum number of patients who need an ICU bed and respiratory were estimated as 3.49 [3.16, 3.71] million per day.

Results of policy simulation for SC and VEC are presented in Table 2: SC alone can reduce the total number of symptomatic patients by five million (7%); VEC can reduce it by six million (8%). Performing SC and VEC simultaneously can reduce patients by 12 million (17%). The 95% CI of the total numbers of patients with SC and VEC over the base case were [0.910, 0.948], [0.869, 0.939], and [0.781, 0.833]. Therefore, all interventions were found to reduce the number of patients significantly.

**Table 2:** Results of estimation of patients under interventions of four types

|  | No intervention |  |  | School closure |  |  |
| --- | --- | --- | --- | --- | --- | --- |
|  | Median | 95%CI |  | Median | 95%CI |  |
|  |  | Lower | Upper |  | Lower | Upper |
| Total number of patients | 78.6 | 69.4 | 83.3 | 73.2 | 65.6 | 76.0 |
| Maximum number of patients | 2.25 | 1.85 | 2.58 | 1.84 | 1.57 | 1.96 |
| Maximum number of inpatients | 12.9 | 11.7 | 13.8 | 11.3 | 10.4 | 11.8 |
| Maximum number of ICU patients | 3.49 | 3.16 | 3.71 | 3.05 | 2.79 | 3.18 |
|  | Voluntary Event Cancellation |  |  | Both |  |  |
|  | Median | 95%CI |  | Median | 95%CI |  |
|  |  | Lower | Upper |  | Lower | Upper |
| Total number of patients | 72.6 | 60.4 | 78.3 | 64.7 | 54.3 | 68.4 |
| Maximum number of patients | 1.73 | 1.19 | 2.15 | 1.23 | 0.91 | 1.36 |
| Maximum number of inpatients | 0.48 | 8.23 | 111.87 | 8.12 | 6.56 | 8.75 |
| Maximum number of ICU patients | 2.81 | 2.21 | 3.18 | 2.18 | 1.76 | 2.35 |

Note: This table presents simulation results of the total number of patients based on the base case (no intervention), school closure, voluntary event cancellation and both through 10,000 iterations of bootstrapping.

## Discussion

We applied a simple SIR model with three age classes including asymptomatic cases and assuming some proportion of children as protected. An earlier study [11–13] estimated R_0_ for COVID-19 as 2.24–3.58 in Wuhan. Our obtained R_0_ was similar. However, we estimated the protection level among children as 40%, even though it had a wide value for 95%CI. Nevertheless, this study was the first to estimate it and to confirm rejection of the null hypotheses that the protection level was very low around 0.1 or very high around 0.9.

We also demonstrated that SC and VEC have significant effects on reducing the total number of patients. Performed simultaneously, SC and VEC can reduce the total number of patients by 18%. However, one must be reminded that effects of SC and VEC on contact patterns and frequencies were based on assumptions. No evidence was found to verify those assumptions. Therefore, the results must be interpreted as representing one possibility.

## Conclusion

Some significant cross protection was found among children for COVID-19. Moreover, SC and VEC were found to have significant effects on reducing the total number of patients. Actually, on March 10, SC and VEC were extended until March 19. We hope that the present study contributes to government decision-making for SC and VEC. The present study represents the authors’ opinion. It does not reflect any stance of our affiliation.

## Data Availability

Japan Ministry of Health, Labour and Welfare. Press Releases of Domestic Situation (in Japanese)

https://www.mhlw.go.jp/stf/seisakunitsuite/bunya/0000121431_00086.html

## Reference

1. Japan Ministry of Health, Labour and Welfare. Press Releases of Domestic Situation (in Japanese) https://www.mhlw.go.jp/stf/seisakunitsuite/bunya/0000121431_00086.html [accessed on 17 March, 2020]

2. Lee PI, Hu YL, Chen PY, Huang YC, Hsueh PR. Lee PI, et al. Are children less susceptible to COVID-19? J Microbiol Immunol Infect. 2020 Feb 25:S1684-1182;30039-6.

3. Japan Ministry of Health, Labour and Welfare. Press Releases. https://www.mhlw.go.jp/stf/houdou/houdou_list_202001.html. (in Japanese) [accessed on 18 March, 2020]

4. Ohkusa Y, Sugawara T, Taniguchi K, Okabe N. Real-time estimation and prediction for pandemic A/H1N1(2009) in Japan. J Infect Chemother. 2011;17:468–72.

5. Ferguson NM, Cummings DA, Cauchemez S, Fraser C, Riley S, Meeyai A, et al.: Strategies for containing an emerging influenza pandemic in Southeast Asia. Nature 2005; 437:209–214.

6. Longini IM Jr, Nizam A, Xu S, Ungchusak K, Hanshaoworakul W, Cummings DA, Halloran ME: Containing Pandemic Influenza at the Source. Science 2005; 309:1083–1087.

7. Germann TC, Kadau L, Longini IM Jr, Macken CA: Mitigation strategies for pandemic influenza in the United States. Proc Natl Acad Sci USA 2006; 103: 5935–5940.

8. Ferguson NM, Cummings DA, Fraser C, Cajka JC, Cooley PC, Burke DS: Strategies for mitigating an influenza pandemic. Nature 2006; 442: 448–452.

9. Ohkusa Y, Sugawara T: Simulation Model of Pandemic influenza in the Whole of Japan, Journal of Japanese Infectious Disease 2009; 62:98–106.

10. Ibuka Y, Ohkusa Y, Sugawara T, Chapman GB, Yamin D, Atkins KE, Taniguchi K, Okabe N, Galvani AP. Social contacts, vaccination decisions and influenza in Japan. J Epidemiol Community Health 2016;70:162–7.

11. Zhao S, Lin Q, Ran J, Musa SS, Yang G, Wang W, Lou Y, Gao D, Yang L, He D, Wang M. Preliminary Estimation of the Basic Reproduction Number of Novel Coronavirus (2019-nCoV) in China, From 2019 to 2020: A Data-Driven Analysis in the Early Phase of the Outbreak. Int J Infect Dis 2020 [Online ahead of print]

12. Liu Y, Gayle AA, Wilder-Smith A, Rockly J. The reproductive number of COVID-19 is higher compared to SARS coronavirus. J Travel Med. 2020.DOI: 10.1093/jtm/taaa021

13. Lai C, Shih T, Ko W, Tang H, Hsueh P. Severe Acute Respiratory Syndrome Coronavirus 2 (SARS-CoV-2) and Coronavirus disease-2019 (COVID-19): The Epidemic and the Challenges. Int J Antimicrob Agents. DOI:10.1016/j.ijantimicag.2020.105924

